# Associations between Urinary Pesticide Metabolites and Serum Inflammatory Biomarkers in Adolescents Living in an Agricultural Region

**DOI:** 10.1101/2024.10.22.24315869

**Authors:** Mohammed N. Hussari, Briana N.C. Chronister, Kun Yang, Xin Tu, Danilo Martinez, Rajendra P Parajuli, Jose Suarez-Torres, Dana Boyd Barr, Suzi Hong, Jose R. Suarez-Lopez

**Affiliations:** University of California San Diego (UCSD) School of Medicine, 9500 Gillman Dr, CA 92093, USA; Herbert Wertheim School of Public Health and Human Longevity Science, University of California San Diego (UCSD), 9500 Gillman Dr. MC: 0725, 92093-0725 CA, USA; School of Public Health, San Diego State University, San Diego, California, USA; Fundación Cimas del Ecuador (CIMAS), De los Olivos E14-226 y, Quito 170124, Ecuador; Rollins School of Public Health, Emory University, 1518 Clifton Road, NE, Atlanta, GA 30322, USA; Department of Psychiatry, School of Medicine, University of California San Diego

**Keywords:** Inflammation, Pesticides, Agriculture, Ecuador, Cytokines, Adolescents

## Abstract

**Background:** In-vitro and in-vivo studies have shown evidence for the immuno-modulatory properties of different pesticides. However, few epidemiological studies on inflammation and pesticide exposure exist, with none in children and adolescents. Associations between pesticide metabolites in urine and inflammatory biomarkers in serum were evaluated among children and adolescents (n=512) of rural Ecuador as part of the ESPINA study

**Methods:** Seventeen urinary biomarkers of insecticide, herbicide and N, N-diethyl-meta-toluamide (DEET) insect repellent exposure were measured. Among them, acetamiprid-N-desmethyl [AND], 3,5,6-Trichloro-2-pyridinol [TCPy], *para*-nitrophenol [PNP], malathion dicarboxylic acid [MDA], 3-phenoxybenzoic acid [3-PBA], 2,4-Dichlorophenoxyacetic acid [2,4-D], glyphosate and 3-(diethylcarbamoyl) benzoic acid [DCBA] were included in analyses as they were detected in >30% of participants. Serum analysis included c-reactive protein (CRP), interleukin-6 (IL-6), tumor necrosis factor-⍺ (TNF-⍺), soluble intercellular adhesion molecule-1 (sICAM-1), soluble vascular cell adhesion molecule-1 (sVCAM-1), serum amyloid A (SAA), and soluble CD14 (sCD14). Associations were evaluated by generalized estimating equations (GEE) and partial least squares (PLS) regression, adjusting for demographic, anthropometric and socioeconomic variables.

**Results:** Positive quadratic associations were found between 2,4-D and CRP (β^2^=0.13, [0.00, 0.27]), IL-6 (β^2^=0.10, [0.04, 0.15]), SAA (β^2^=0.13, [0.01, 0.30]), sICAM-1 (β^2^=53.25, [27.26, 79.24]) and sVCAM-1 (β^2^=61.11, [30.52, 91.70]). The pesticide metabolites PLS composite variable was positively associated with IL-6 (β=0.09, [0.01. 0.17]), SAA (β=0.43, [0.13, 0.17]), sICAM-1 (β=63.52, [9.92, 117.13]), sVCAM-1 (β=69.03, [8.29, 129.76]) and TNF-⍺ (β=0.08, [0.00, 0.16]), and negatively associated with CRP (β=-0.28, [-0.49, -0.08], Figure 1).

**Conclusions:** Our findings demonstrate a novel pesticide/herbicide-inflammation link in adolescents, which may be an underlying mechanism of the health impacts of pesticides/herbicides.

## Introduction

Pesticide use in agriculture has soared in recent decades due to the Green Revolution, which introduced agrochemical use in human farming practices (Hayes and Hansen 2017). The Green Revolution’s technological package increased agricultural yields and profits but came with significant environmental and health consequences (Suárez-Torres et al. 2017). In addition to acute toxicities, there is emerging evidence from human, animal and in-vitro models that chronic, low-level exposure to pesticides can have significant health consequences, including neurocognitive and neurodegenerative disorders such as ADHD, Parkinson’s disease, other pathologies including bladder and hematologic cancers, and changes in cytokine mRNA expression (Henderson et al. 2002; de Souza et al. 2011; Baltazar et al. 2014; Koutros et al. 2016).

Although various studies have demonstrated associations between inflammation status and the use of agrochemicals, many of the studies examining the relationship between inflammation and pesticides are largely confined to in vitro and animal models (Agrawal et al. 2015a; Singh et al. 2016; Patel et al. 2018; Miao et al. 2022; Deng et al. 2022). Only a handful of studies have illustrated the effects of pesticide exposure on inflammation in humans.

Inflammation, particularly chronic systemic inflammation, has been implicated in the pathogenesis of the world’s most disabling and deadly diseases, including cardiovascular disease, chronic kidney disease, diabetes mellitus, non-alcoholic fatty liver disease, neurodegenerative disease and autoimmune disease (Furman et al. 2019).

Growing evidence indicates that human agricultural workers are at increased risk for inflammatory diseases. A meta-analysis of 14 different studies that examined pesticide exposure across various classes found evidence for associations between various pesticides and Parkinson’s disease (Huang et al. 2022), which is believed to involve a neuroinflammatory process in its pathogenesis (Priyadarshi et al. 2001; Tansey et al. 2022; Huang et al. 2022). Similarly, a study of adult male pesticide applicators found that those diagnosed with rheumatoid arthritis (RA) had greater intensity-weighted lifetime use of certain insecticides and herbicides than did those without a confirmed RA diagnosis (Armando et al. 2024). Another study found that participants with recently diagnosed systemic lupus erythematosus (SLE) had higher odds of having ever worked in pesticide mixing (Cooper et al. 2004). Similarly, in a cohort of greenhouse insecticide applicators, the use of neonicotinoids, organophosphates, and pyrethroids was associated with elevated levels of interleukin-22 (IL-22), a cytokine with complex pro- and anti-inflammatory functions. (Fenga et al. 2014). In contrast to these findings, a study found that male pesticide sprayers exposed to the pyrethroid cypermethrin had reduced levels of the proinflammatory cytokines IL-2, IL-8, IL-12 and interferon-gamma (IFN-γ) compared to those of male office workers who served as controls with less pesticide exposure. (Costa et al. 2013).

A limited number of studies have characterized the associations between pesticide exposure and inflammation biomarkers, and none have been found in pediatric populations. Mwanga et al. performed a cross-sectional study involving 113 women residing on farms in South Africa, and 98 women living in neighboring towns to examine the associations between urinary pesticide metabolites and various outcomes, including cytokine profiles for IL-4, IL-5, IL-6, IL-8, IL-10, IL-13 and IL-17, and found a consistent trend of increased cytokines with higher concentrations of various organophosphate and pyrethroid metabolites (Mwanga et al. 2016). The association between use of pesticide mixtures, as reported through questionnaires, and various cytokines was studied in a group of 108 floriculture workers in Mexico, and it was found that the levels of IL-6 and IFN-γ were higher amongst those with greater exposure (Godínez-Pérez et al. 2024). One small study involving 31 farm workers occupationally exposed to pesticides and city-dwelling unexposed controls matched for age and sex found significant increases in IL-6 concentrations among participants categorized as exposed (El-Gameel et al. 2023). Another small study in Brazil compared the cytokine levels of farmworkers with reported occupational pesticide exposure to those of healthy controls and revealed that IL-6 was elevated in the exposed group (Jacobsen-Pereira et al. 2020). Similarly, Da Silva et al. reported that women with breast cancer who reported occupational exposure to pesticides had the worst prognostic features, a shift in helper T-cell (Th) profiles from Th1 to Th2, and a decrease in IL-17A, IL-1B and IL-4 in those with lymph node invasion (da Silva et al. 2022b).

Studies using in vitro and animal models have supported findings from human observational studies, demonstrating that insecticide exposure can upregulate inflammation. This includes upregulation of nuclear factor kappa beta (NF-kβ), tumor necrosis factor alpha (TNF-α), IL-1β, and IL-6 associated with neonicotinoid insecticide exposure (Miao et al. 2022; Deng et al. 2022), as well as elevated pro-inflammatory markers in the substantia nigra and microglia (Agrawal et al. 2015a, b; Hossain et al. 2017). Additionally, pyrethroid exposure has been linked to increased reactive oxygen species (ROS) production, while organophosphate insecticide exposure has been associated with the upregulation of IL-8 and TNF-α and the downregulation of the anti-inflammatory cytokine IL-10 (Schäfer et al. 2013; Proskocil et al. 2019). Organophosphates, and to a lesser extent the insect repellent N,N-Diethyl-3-methylbenzamide (DEET), are known to inhibit acetylcholinesterase (AChE) activity (Corbel et al. 2009), and alterations in the cholinergic system due to pesticide exposure can trigger inflammatory responses and related pathophysiological changes or lead to reduced regulation of inflammation via the cholinergic anti-inflammatory pathway (Mokarizadeh et al. 2015).

Herbicides represent another major class of pesticides, and glyphosate and 2,4-D are the two most used herbicides worldwide. Animal and in vitro studies have found 2,4-D exposure to be associated with increased inflammasome activation and elevated caspase-1, IL-1β, IL-18, and p62 protein expression (Zhou et al. 2022), and reduced glutathione stores in human erythrocytes, suggesting that it is an oxidative stressor (Bukowska 2003). Glyphosate was found to upregulate the expression of IL-1β, TNF-α, IL-6, and C-reactive protein (CRP) in liver tissue samples compared to controls (Pandey et al. 2019), and was associated with increased IL-33, TSLP, IL-13, and IL-5, and increased numbers of eosinophils and neutrophils in lung tissue (Kumar et al. 2014). Glyphosate may disturb mitochondrial architecture and function, inducing ROS-mediated inflammation (Qi et al. 2023).

Given the limited research on inflammation in humans as a function of pesticide exposure, our study seeks to better understand whether urinary pesticide metabolites, representing exposure to varying classes of pesticides, are associated with human inflammation markers in adolescents living in agricultural settings in Ecuador. It is important to understand how inflammation status is affected by pesticide exposure given the emerging evidence for inflammation-mediated pathogenesis of various diseases (Furman et al. 2019). We hypothesized that insecticide and herbicide urinary metabolites would positively associate with serum inflammatory biomarkers.

## Materials and Methods

### Study Population

The study of Secondary Exposures to Pesticides among Children and Adolescents (ESPINA) is a prospective cohort study focused on understanding the effects of pesticide exposure on the development of children and adolescents residing in the agricultural county of Pedro Moncayo, Pichincha Province, Ecuador. The ESPINA study was established in 2008 and examined 313 boys and girls aged 4-9 years. Participants were recruited mostly through community announcements for their participation in the “2004 Survey of Access and Demand of Health Services in Pedro Moncayo County,” a representative survey of Pedro Moncayo County collected by Fundación Cimas del Ecuador in conjunction with the communities of Pedro Moncayo County. Details about participant recruitment in 2008 have been described previously (Suarez-Lopez et al. 2013). In 2016, the cohort was expanded, and two examinations were conducted. We examined a total of 554 participants in 2016: 330 participants in April and 535 in July-October, with 311 participants examined during both time periods. Participants in 2016 (Suarez-Lopez et al. 2020) were 12-17 years of age and included 238 participants examined in 2008 and 316 new volunteers (Suárez-López et al. 2020). The System of Local and Community Information (SILC) developed by Fundación Cimas del Ecuador was used to recruit new participants in 2016. The SILC included information from the 2016 Community Health Survey of Pedro Moncayo County (formerly the Survey of Access and Demand of Health Services in Pedro Moncayo). None of the participants reported working in agriculture in either 2008 or 2016. Additional details of participant recruitment in 2016 have been published (da Silva et al. 2022a).

To maximize the number of participants included in our analyses, we imputed missing data for parental education in 2016 (n=10), as well as residential distance to the nearest flower plantation in 2008 (n=3). Among the children who were examined in 2008 but were missing parental education data in 2016 (n=5), data for parental education were imputed using the 2008 data for maternal and paternal education. For participants with missing paternal and maternal education data in 2008 and missing parental education in 2016 (n=5), a random imputation was conducted for each variable based on a normal distribution of the variable during the respective examination period. Since a small number of participants reported being White (n=4), we grouped these 6 participants in the mestizo (mix of White and Indigenous) category to improve model stability when adjusting for ethnicity.

The present analysis includes 512 participants examined in the July-October 2016 who had all covariates of interest: 8 observations were excluded due to missing inflammatory marker data and another 15 due to missing covariate data.

The 2016 examination of the ESPINA study was approved by the Institutional Review Boards of the University of California San Diego (UCSD), Universidad San Francisco de Quito and the Ministry of Public Health of Ecuador and endorsed by the Commonwealth of Rural Parishes of Pedro Moncayo County. We obtained informed consent from adult participants (aged 18 years or older) and parents, as well as parental permission of participation and informed assent of child participants.

## Data Collection

### Examination and Survey

In 2016, children were examined in 7 schools when they were out of session between July and October. The examiners were unaware of the participants’ potential exposures. Home interviews with parents and adult residents were conducted to gather socioeconomic and demographic information. Participant ethnicity was categorized into 2 groups: 114 Indigenous and 407 Mestizo participants.

### Anthropometric measures

Participant height was assessed using a height board following recommended guidelines, and weight was measured using a digital scale (Tanita model 0108 MC; Tanita Corporation of America, Arlington Heights, IL, USA). We calculated z-scores for height-for-age (Z-height-for-age) and body mass index-for-age (Z-BMI-for-age) using the World Health Organization growth standards (World Health Organization 2006).

### Biospecimen analysis

All biospecimens were collected on the same day, including urine and venous blood.

### Inflammation markers

Venipuncture of the median cubital or cephalic veins in the arm was conducted by a phlebotomist following standard guidelines without fasting (Clinical and Laboratory Standards Institute 2007). Blood samples were centrifuged and aliquoted on site and transported on dry ice to Quito, approximately 45-55 km from the various blood collection sites across Pedro Moncayo, for storage at -70°C. Samples were then shipped to the University of California San Diego (UCSD) at -20°F using an overnight courier. Samples were stored at -80°C at UCSD until they were analyzed for inflammation marker quantification at the UCSD Integrative Health Mind-Body Biomarker Core. Multiplex assay kits were used to measure plasma levels of pro-inflammatory markers including TNF-α, IL-6, CRP, soluble intercellular adhesion molecule-1 (sICAM-1), serum amyloid A (SAA) and soluble vascular adhesion molecule-1 (sVCAM-1), using the 2400 SECTOR Imager reader (Meso Scale Discovery, Rockville, Maryland). Levels of soluble cluster of differentiation 14 (sCD-14), a pro-inflammatory marker, indicating monocyte activation, were measured using quantikine ELISA kits using R&D Systems (Minneapolis, MN). The intra- and inter-assay coefficients of variation of those markers were <10%. The Pearson’s correlation coefficients between paired inflammation markers were calculated and tested as additional information (Table S1).

### Urinary pesticide biomarkers

#### Overview

Seventeen urinary biomarkers of pesticide exposure were measured, including six neonicotinoid metabolites (acetamiprid [ACET], acetamiprid-N-desmethyl [AND], clothianidin [CLOT], imidacloprid [IMID] and thiacloprid [THIA]), four organophosphates (2-isopropyl-4-methyl-6-hydroxypyrimidine [IMPy], 3,5,6-Trichloro-2-pyridinol [TCPy], *para*-nitrophenol [PNP], and malathion dicarboxylic acid [MDA]), three pyrethroid metabolites (3-phenoxybenzoic acid [3-PBA], 4-fluoro-3-phenoxybenzoic acid [4F-3PBA] and trans-3-(2,2-Dichlorovinyl)-2,2-dimethylcyclopropane carboxylic acid [*trans-DCCA]*), two herbicides (2,4-Dichlorophenoxyacetic acid [2,4-D] and glyphosate) and two DEET metabolites (3-(diethylcarbamoyl) benzoic acid [DCBA] and 3-(ethylcarbamoyl) benzoic acid [ECBA]). Urine samples were shipped overnight at -20° C from UCSD to the National Center for Environmental Health, Division of Laboratory Sciences of the CDC (Atlanta, GA) for quantification of all organophosphates, neonicotinoids, pyrethroids, DEET and 2,4-D biomarkers, and to the Laboratory for Exposure Assessment and Development in Environmental Research at Emory University (Atlanta, GA) for quantification of creatinine and glyphosate. Quality control/quality assurance protocols were followed to ensure data accuracy and reliability of the analytical measurements. All the study samples were re-extracted if quality control failed the statistical evaluation (Baker et al. 2019).

#### Sample collection and preparation

Urinary concentrations of creatinine and pesticide biomarkers were measured in samples collected upon awakening. Participants brought the urine samples to the examination site in the morning where they were aliquoted and frozen at -20° C. At the end of each day, samples were transported to Quito for storage at -80° C. Samples were then shipped frozen overnight to UCSD for long-term storage at -80° C.

#### Urinary organophosphate, pyrethroid, and 2,4-D biomarker quantification

Organophosphate and pyrethroid metabolites and 2,4-D were quantified using liquid chromatography coupled with tandem mass spectrometry and isotope dilution (Davis et al. 2013; Baker et al. 2019). The limit of detection (LOD) was 0.1 μg/L for TCPy, PNP, IMPy, 3-PBA, and 4F-3PBA; 0.15 μg/L for 2,4-D; 0.5 μg/L for MDA; and 0.6 μg/L for *trans*-DCCA. Targeted metabolites were quantified from enzymatic hydrolysis of 0.5 or 1.0 mL of urine (metabolite dependent) and online solid phase extraction to release, extract and concentrate the target biomarkers, followed reversed-phase high-performance liquid chromatography-tandem mass spectrometry (HPLC-MS/MS) using electrospray ionization (ESI) (Davis et al. 2013; Baker et al. 2019). The LOD was 0.03 μg/L for THIA, 0.2 μg/L for AND and CLOT, 0.3 μg/L for ACET, and 0.4 μg/L for IMID and OHIM.

#### Urinary glyphosate quantification

Urine aliquots (250 μL) were spiked with isotopically labeled glyphosate, diluted to 1 mL with doubly deionized water, and extracted using a C18 solid phase extraction (SPE). Glyphosate was derivatized to create its heptafluorobutyl analogue then concentrated for analysis. To measure urinary glyphosate, all urine samples (aliquots) were randomized using a Fisher-Yates shuffling algorithm prior to analysis to reduce potential batch effects (Fisher and Yates 1948; Knuth 1960). Concentrated extracts were analyzed using gas chromatography-mass spectrometry using electron impact ionization in the multiple ion monitoring mode. The LOD was 0.25 μg/L with a relative standard deviation (RSD) of 3%.

#### Urinary Neonicotinoid and DEET biomarker quantification

The neonicotinoid and DEET metabolite quantification analytical method is based on enzymatic hydrolysis of 0.2 mL of urine and online solid phase extraction to release, extract and concentrate the target biomarkers, followed reversed-phase high-performance liquid chromatography-tandem mass spectrometry (HPLC-MS/MS) using electrospray ionization (ESI). Additional details have been described previously (Baker et al. 2019). The LOD for ECBA and DCBA was 0.2 μg/L. The precision of the measurements, expressed as the percent RSD of multiple measures of two urine-based quality control (QC) materials, was below 6%, depending on the biomarker and concentration.

#### Imputation for values below the LOD

After excluding any biomarkers with a detection rate below 30%, nine were included in our analysis: TCPy, PNP, MDA, 3-PBA, AND, 2,4-D, glyphosate and DCBA. We used multiple imputation to estimate metabolite concentrations for individuals who had pesticide metabolite concentrations below the LOD. The multiple imputation method was built as a log-logistic regression model. The initial model was adjusted for: age, gender, race, BMI-for-age z-score (z-BMI-for-age), monthly family income and parental education. Predicted concentrations were then rescaled between 0 and the LOD and imputed for observations below the level of detection. For the pesticide summary score calculation (see below) a variable that is only used to describe participant characteristics across a pesticide exposure construct in Table 1, observations below the LOD were imputed with a constant (LOD divided by the square root of two). This constant imputation method was also applied when we create the composite variables of pesticide exposure in PLS regression.

**Table 1.** Characteristics of study participants.

| **Table 1.** Characteristics of study participants |  |  |  |  |  |
| --- | --- | --- | --- | --- | --- |
|  | **All** | **Tertile of Pesticide Summary Score** |  |  |  |
| **N** | **507** | **T1**<br>**168** | **T2**<br>**167** | **T3**<br>**168** | **p-trend** |
| **Range** |  | **0.28, 1.26** | **1.27, 1.77** | **1.78, 5.02** |  |
| Age (years) | 14.5 ± 1.77 | 14.4 ± 1.75 | 14.4 ± 1.78 | 14.6 ± 1.77 | *p*=0.548 |
| Gender |  |  |  |  |  |
| Male (%) | 50% | 52% | 55% | 44% | *p*=0.158 |
| Female (%) | 50% | 48% | 45% | 56% |  |
| Race |  |  |  |  |  |
| Indigenous (%) | 22% | 27% | 21% | 19% | *p*=0.106<sup>c</sup> |
| Mestizo (%) | 78% | 73% | 79% | 81% |  |
| BMI-for-age Z-score (SD) | 0.384 ± 0.835 | 0.448 ± 0.753 | 0.364 ± 0.860 | 0.341 ± 0.888 | *p*=0.453 |
| Family's Monthly Income (USD) | 500 (350) | 460 (335) | 500 (325) | 600 (374) | *p*=0.334 |
| Average Parental Education (years) | 7.00 (4.50) | 7.00 (4.50) | 7.00 (3.50) | 7.50 (4.50) | *p*=0.316 |
| Presence of Smokers at Home |  |  |  |  |  |
| No | 77% | 81% | 78% | 72% | *p*=0.080 |
| Yes | 23% | 19% | 22% | 28% |  |
| TCPy (ug/L) | 2.69 (2.89) | 1.44 (1.13) | 2.78 (2.55) | 4.39 (3.97) | *p*<0.001 |
| PNP (ug/L) | 0.505 (0.371) | 0.350 (0.194) | 0.512 (0.268) | 0.817 (0.626) | *p*<0.001 |
| MDA (ug/L) | 0.819 (0.188) | 0.771 (0.139) | 0.813 (0.134) | 0.876 (0.213) | *p*=0.107 |
| DCBA (ug/L) | 0.769 (2.54) | 0.540 (0.751) | 0.798 (3.54) | 0.833 (4.46) | *p*=0.694 |
| 3-PBA (ug/L) | 0.428 (0.504) | 0.258 (0.172) | 0.407 (0.317) | 0.822 (1.12) | *p*<0.001 |
| AND (ug/L) | 0.573 (0.766) | 0.521 (0.429) | 0.573 (0.867) | 0.588 (1.00) | *p*=0.069 |
| 2,4-D (ug/L) | 0.303 (0.266) | 0.213 (0.106) | 0.266 (0.192) | 0.423 (0.352) | *p*<0.001 |
| Glyphosate (ug/L) | 0.870 (1.38) | 0.390 (0.516) | 0.898 (1.08) | 1.85 (2.41) | *p*<0.001 |
| AChE (U/mL) | 3.70 ± 0.553 | 3.69 ± 0.573 | 3.72 ± 0.520 | 3.71 ± 0.568 | *p*=0.635 |
| sCD14 (ng/mL) | 1590 ± 392 | 1620 ± 384 | 1570 ± 384 | 1580 ± 408 | *p*=0.617 |
| CRP (µg/mL) | 1.39 ± 1.66 | 1.48 ± 1.66 | 1.37 ± 1.70 | 1.33 ± 1.61 | *p*=0.427 |
| IL-6 (pg/mL) | 1.18 ± 0.684 | 1.15 ± 0.610 | 1.18 ± 0.714 | 1.20 ± 0.725 | *p*=0.084 |
| SAA (µg/mL) | 2.01 ± 2.05 | 2.06 ± 1.99 | 1.88 ± 1.99 | 2.10 ± 2.17 | *p*=0.500 |
| sICAM-1 (ng/mL) | 755 ± 467 | 752 ± 431 | 723 ± 483 | 788 ± 484 | *p*=0.157 |
| sVCAM-1 (ng/mL) | 975 ± 548 | 966 ± 510 | 924 ± 554 | 1030 ± 577 | *p*=0.031 |
| TNF-α (pg/mL) | 2.11 ± 0.609 | 2.13 ± 0.590 | 2.07 ± 0.498 | 2.13 ± 0.719 | *p*=0.663 |

Values displayed mean ± standard deviation, percent or median and interquartile range (IQR).
Participant demographics did not differ significantly across summarized pesticide score tertile.
2,4-D= 2,4-Dichlorophenoxyacetic acid; AChE=acetylcholinesterase; AND=acetamiprid-N-desmethyl; TCPy= 3,5,6-Trichloro-2-pyridinol; DCBA=3-(diethylcarbamoyl) benzoic acid; MDA= Malathion dicarboxylic acid; 3-PBA=3-phenoxybenzoic acid; PNP= para-Nitrophenol.

### Urinary Creatinine quantification

To account for urinary dilution, the urinary creatinine concentration was quantified using HPLC-MS/MS with ESI. A 10 μL aliquot of urine was diluted prior to analysis (Kwon et al. 2012). No further sample preparation was performed prior to analysis. The LOD was 5 mg/dL, with an RSD of 7%.

### AChE activity and hemoglobin concentration

AChE activity and hemoglobin concentration were measured in fresh finger-stick blood samples using the EQM-Test-mate ChE Cholinesterase Test System 400 during the examination following recommended procedures (EQM Research Inc., Cincinnati, OH, USA). Lower concentrations of AChE activity reflect greater exposure to cholinesterase inhibitor pesticides (Skomal et al. 2022).

### Statistical analysis

#### Pesticide Summary Score Calculation

The pesticide summary score represents the amount of pesticide exposure that a participant was found to have. This score was only used in Table 1 to describe participant characteristics across a single construct of urinary pesticide concentrations. The summarized pesticide score was calculated using the following steps. One was added to the variable and was natural-log transformed. The log-transformed concentrations were divided by the group’s standard deviation, and then averaged with all pesticide metabolites, a variation of a method used in previous work (Suarez-Lopez et al. 2015). The metabolites that were included in the summarized score were TCPy, PNP, MDA, 3-PBA, AND, DCBA, 2,4-D and glyphosate.

Descriptive statistics were calculated for the overall cohort, and across tertiles of the summarized pesticide score. We calculated column percents for count data, and the means and standard deviations (SDs) for all normally distributed continuous variables. To determine whether there was a linear trend for participant characteristics across pesticide metabolite concentrations, we calculated a p-value for trend (p-trend) by running a linear regression using the summarized pesticide score as the independent variable, and the characteristic as the dependent variable. If the p<0.05, then there was a linear trend. All pesticide concentration variables were naturally log-transformed because of non-normal distribution.

#### Independent associations of pesticide and DEET biomarkers with inflammation markers

Associations between log-transformed pesticide metabolites and inflammatory marker levels were estimated using linear regression with GEE, adjusting for covariates identified a priori. The covariates included were age, gender, race, BMI-for-age z-score, height-for-age z-score, parental education level, urinary creatinine, smokers at home (yes/no) based on research in the literature that indicates these variables may have associations with various inflammatory markers (Singer et al. 2014; Artero et al. 2014; Conklin et al. 2019; Klevebro et al. 2021; Horton 2023). When the independent variable was AChE, the models also adjusted for hemoglobin concentrations as the AChE is the erythrocytic concentration of the enzyme. In linear models, pesticide metabolite and AChE values were log-transformed and estimates of coefficients were scaled to show the difference in inflammatory biomarkers given a 50% increase in the metabolites or AChE.

To investigate if the individual pesticide metabolites had a curvilinear association with each inflammatory biomarker, we tested the significance of a quadratic term (e.g., ln_TCPy*ln_TCPy) into the regression model. Conversion per a 50% increase in metabolites, as described above, was not necessary for quadratic models, as the interpretation of the coefficients mostly focuses on the shape of the association. A curvilinear relationship was identified if the squared term had a p<0.05. To visualize the adjusted associations between individual pesticide metabolites and individual inflammatory biomarkers, adjusted least squares means of inflammatory biomarkers were calculated for up to 1000 ranks for each metabolite and were plotted. Based on the covariate-adjusted data for the ranked variables, LOESS curves were used to visualize the adjusted associations with 95% CI.

#### Associations of pesticide and DEET biomarker mixtures with inflammation markers

Given the multiple exposure variables, partial-least-squares (PLS) regression was used to create a composite exposure variable. PLS was chosen instead of principal component analysis (PCA) or factor analysis (FA), because it is more effective when composite variables are used as independent variables in a regression analysis (Liu et al. 2022). PLS regression is similar to PCA and FA in that it is a variable reduction methodology and creates a composite variable that is a linear combination of the original variables. The new composite variables are orthogonal and therefore uncorrelated. Unlike PCA, which orders its composite variables based on variance, with the first composite variable explaining the most variance, PLS orders the composite variables such that the first composite is the one that is most correlated with the outcome (Liu et al. 2022). Moreover, PLS is also used in cases where the explanatory variables are collinear and therefore problematic when used in a multiple linear regression, or when there are multiple explanatory variables with possible additive or synergistic effects.

The composite variable is a linear combination of the explanatory variables, and the loadings represent the direction and magnitude of the individual explanatory variables’ contributions to the composite variable. In our analysis, the composite variable included five standardized pesticide biomarkers (glyphosate, TCPy, 2,4-D, 3-PBA and PNP), each of which could contribute positively or negatively to the composite-dependent variable association. To include as many participants as possible, imputation on 2,4-D and 3-PBA was considered. The multiple imputation was not appropriate because it would introduce more variables which could make the composite variables more complicated. Similar to the calculation of our pesticide summary score, we applied the constant imputation method instead. A positive vector loading for a given metabolite indicates that the metabolite is associated with the dependent variable in the same direction as the composite variable, and vice versa for negative loadings. Winsorization was used to reduce the effect of outliers in the outcome variables on the PLS analysis.

Particularly for this study, a residualization procedure was adopted as adjustment for covariates, which means the outcome variable corresponding to each inflammatory biomarker was the residual from a linear model fitted with the original outcome and the covariates. Negative R-squared values were set to zero. Selection of which composite variables to include in the regression model was based on p-value, effect size and the adjusted R-squared.

## Results

### Participant Characteristics

Participant characteristics for the overall cohort (n=507) stratified by tertiles of the pesticide summary score can be found in Table 1. Overall, the mean age of participants was 14.5 years (standard deviation [SD]=1.77 years), 50% of the participants were male, and the racial composition was 22% indigenous and 78% mestizo. None of the participant sociodemographic characteristics differed significantly across the pesticide summary score. As expected, many of the pesticide metabolites differed significantly across the pesticide summary score with the exception of for MDA and AND.

### Associations between inflammatory biomarkers amongst themselves

The majority of inflammatory biomarkers exhibited few correlations amongst themselves (Table S 1). Levels of CRP showed significant positive associations with the levels of SAA, sICAM, sVCAM and TNF-*α*.

### Associations between pesticide and DEET biomarkers and inflammatory biomarkers

Independent linear associations were analyzed for each pesticide metabolite and the inflammatory biomarkers using GEE models. No significant associations were found between metabolites and the inflammation biomarkers in the linear models (Table S2).

Analyses assessing curvilinear (quadratic) associations between pesticide biomarkers and inflammation are presented in Table 2. Notably, we found significant and consistently positive curvilinear associations of 2,4-D with CRP (β^2^= 0.13, [0.00, 0.27]), IL-6 (β^2^= 0.10, [0.04, 0.15]), SAA (β^2^= 0.13, [0.01, 0.30]), sICAM-1 (β^2^=53.25, [27.26, 79.24]) and sVCAM-1 (β^2^= 61.11, [30.52, 91.70]), where β represents percent difference in inflammatory marker concentration for a 10% increase in urinary metabolite concentration (Table 2, Figure 1). Other pesticide and DEET metabolites showed some evidence of curvilinear associations, although the associations were not consistent. Statistically significant negative curvilinear associations were observed for the associations of TCPy with CRP, DCBA with TNF-α, and glyphosate with sCD14; statistically significant positive curvilinear associations were observed for the associations of glyphosate with sVCAM-1 and MDA with TNF-α.

**Table 2.** Independent quadratic associations between biomarkers of pesticide exposure and inflammation N= 507.

| Exposure Biomarkers | Inflammatory Biomarkers |  |  |  |  |  |  |  |
| --- | --- | --- | --- | --- | --- | --- | --- | --- |
| | CRP (ug/ml) | | IL-6 (pg/ml) | | SAA (ug/ml) | | TNF-α (pg/ml) | |
| | $\beta$ (95% CI) | $\beta^2$ (95% CI) | $\beta$ (95% CI) | $\beta^2$ (95% CI) | $\beta$ (95% CI) | $\beta^2$ (95% CI) | $\beta$ (95% CI) | $\beta^2$ (95% CI) |
| **2,4-D** (ug/L) | 0.14<br>[-0.09, 0.37] | **0.13**<br>[**0.00, 0.27**] | 0.08<br>[-0.02, 0.18] | **0.10**<br>[**0.04, 0.15**] | 0.21<br>[-0.08, 0.50] | **0.16**<br>[**0.01, 0.30**] | 0.05<br>[-0.04, 0.14] | 0.00<br>[-0.03, 0.04] |
| **Glyphosate** (ug/L) | -0.05<br>[-0.20, 0.10] | 0.01<br>[-0.02, 0.05] | 0.02<br>[-0.05, 0.10] | 0.00<br>[-0.01, 0.02] | 0.01<br>[-0.20, 0.21] | 0.02<br>[-0.02, 0.05] | 0.03<br>[-0.02, 0.09] | 0.00<br>[-0.01, 0.01] |
| **AND** (ug/L) | 0.04<br>[-0.21, 0.30] | -0.07<br>[-0.24, 0.11] | 0.06<br>[-0.04, 0.16] | -0.03<br>[-0.09, 0.04] | -0.08<br>[-0.41, 0.26] | -0.03<br>[-0.24, 0.18] | -0.11<br>[-0.19, -0.04] | -0.01<br>[-0.08, 0.06] |
| **TCPy** (ug/L) | 0.19<br>[-0.11, 0.49] | **-0.15**<br>[**-0.25, -0.04**] | 0.01<br>[-0.14, 0.16] | -0.01<br>[-0.07, 0.04] | -0.17<br>[-0.60, 0.26] | 0.04<br>[-0.15, 0.22] | 0.04<br>[-0.07, 0.15] | -0.03<br>[-0.07, 0.02] |
| **3-PBA** (ug/L) | -0.09<br>[-0.28, 0.10] | 0.02<br>[-0.10, 0.13] | 0.06<br>[-0.04, 0.16] | 0.02<br>[-0.03, 0.07] | 0.18<br>[-0.12, 0.49] | 0.11<br>[-0.08, 0.30] | 0.02<br>[-0.04, 0.08] | 0.01<br>[-0.03, 0.05] |
| **MDA** (ug/L) | 0.27<br>[-0.51, 1.05] | -0.28<br>[-0.62, 0.05] | -0.22<br>[-0.46, 0.02] | 0.06<br>[-0.05, 0.17] | -0.45<br>[-1.27, 0.36] | 0.01<br>[-0.34, 0.37] | -0.37<br>[-0.66, -0.07] | **0.15**<br>[**0.02, 0.28**] |
| **PNP** (ug/L) | -0.08<br>[-0.56, 0.39] | -0.10<br>[-0.37, 0.18] | -0.06<br>[-0.23, 0.12] | -0.03<br>[-0.15, 0.09] | -0.18<br>[-0.75, 0.40] | -0.06<br>[-0.40, 0.27] | 0.04<br>[-0.10, 0.19] | -0.03<br>[-0.11, 0.06] |
| **DCBA** (ug/L) | -0.05<br>[-0.20, 0.11] | -0.01<br>[-0.05, 0.02] | 0.03<br>[-0.04, 0.09] | -0.01<br>[-0.02, 0.01] | -0.07<br>[-0.26, 0.12] | -0.01<br>[-0.06, 0.04] | 0.05<br>[-0.00, 0.10] | **-0.01**<br>[**-0.03, -0.00**] |
| **AChE** (U/mL) | -8.16<br>[-17.27, 0.95] | 2.93<br>[-0.66, 6.51] | 1.01<br>[-2.33, 4.35] | -0.49<br>[-1.82, 0.84] | -2.59<br>[-14.22, 9.04] | 0.93<br>[-3.70, 5.56] | 0.51<br>[-2.04, 3.06] | -0.20<br>[-1.24, 0.84] |

**Table 2 (continued).** Independent quadratic associations between biomarkers of pesticide exposure and inflammation
| Exposure Biomarkers | Inflammatory Biomarkers |  |  |  |  |  |
| --- | --- | --- | --- | --- | --- | --- |
|  | CD14 (ng/ml) |  | sICAM-1 (ng/ml) |  | sVCAM-1 (ug/ml) |  |
| | $\beta$ (95% CI) | $\beta^2$ (95% CI) | $\beta$ (95% CI) | $\beta^2$ (95% CI) | $\beta$ (95% CI) | $\beta^2$ (95% CI) |
| **2,4-D** (ug/L) | 4.6<br>[-48.3, 57.5] | 5.3<br>[-16.8, 27.4] | 92.4<br>[32.9, 151.9] | **53.25**<br>**[27.3, 79.2]** | 125.5<br>[56.5, 194.5] | **61.1**<br>**[30.5, 91.7]** |
| **Glyphosate** (ug/L) | -1.0<br>[-48.5, 28.6] | **-6.7**<br>**[-12.9, -0.4]** | 30.1<br>[-15.4, 75.6] | **9.6**<br>**[0.4, 18.7]** | 25.1<br>[-25.7, 75.9] | 7.1<br>[-2.0, 16.2] |
| **AND** (ug/L) | 13.1<br>[-53.0, 79.2] | 19.4<br>[-24.2, 62.9] | -19.6<br>[-88.9, 49.7] | -38.8<br>[-82.6, 5.1] | -24.5<br>[-110.1, 61.2] | -39.3<br>[-96.1, 17.5] |
| **TCPy** (ug/L) | 36.9<br>[-40.9, 114.7] | -17.5<br>[-49.7, 14.7] | -20.6<br>[-120.6, 79.5] | 10.8<br>[-30.8, 52.4] | -48.2<br>[-171.0, 74.5] | 19.3<br>[-34.7, 73.3] |
| **3-PBA** (ug/L) | -12.9<br>[-56.9, 31.0] | -1.8<br>[-31.6, 28.7] | 49.4<br>[-21.3, 120.1] | 20.5<br>[-21.7, 62.8] | 64.1<br>[-28.8, 156.9] | 23.1<br>[-33.4, 79.6] |
| **MDA** (ug/L) | -34.3<br>[-237.4, 168.7] | 26.8<br>[-61.4, 114.9] | -108.9<br>[-315.5, 97.8] | 28.2<br>[-59.1, 115.6] | -193.7<br>[-445.5, 58.1] | 73.9<br>[-29.6, 177.3] |
| **PNP** (ug/L) | 29.4<br>[-86.9, 145.7] | 8.1<br>[-60.9, 77.1] | -39.4<br>[-150.1, 71.3] | -35.1<br>[-111.3, 41.1] | -12.1<br>[-160.7, 136.4] | -22.5<br>[-116.0, 71] |
| **DCBA** (ug/L) | 4.6<br>[-34.5, 43.7] | 2.9<br>[-10.3, 16.1] | 1.6<br>[-45.7, 48.9] | -3.2<br>[-16., 9.8] | -1.4<br>[-51.2, 48.4] | -6.8<br>[-20.8, 7.2] |
| **AChE** (U/mL) | 339.8<br>[-1516.1, 2195.7] | -196.4<br>[-949.6, 556.9] | -611.5<br>[-2836.2, 1613.1] | 244.4<br>[-666.9, 1155.7] | -667.6<br>[-2960.5, 1625.4] | 247.5<br>[-695.5, 1190.5] |

2,4-D= 2,4-Dichlorophenoxyacetic acid; AChE=acetylcholinesterase; AND=acetamiprid-N-desmethyl; TCPy= 3,5,6-Trichloro-2-pyridinol; DCBA=3-(diethylcarbamoyl) benzoic acid; MDA= Malathion dicarboxylic acid; 3-PBA=3-phenoxybenzoic acid; PNP= para-Nitrophenol.
Linear GEE model results for each pesticide metabolite and AChE and each inflammatory biomarker.
Presented are the linear and squared $\beta$ coefficients for the quadratic models, where $\beta$ represents the % difference in the inflammatory marker concentration for 1 unit increase in urinary metabolite concentration. Models were adjusted for the following covariates: age, gender, race, BMI-for-age z-score, height-for-age z-score, parental education level, creatinine, smokers at home (yes/no).
Bolded text indicates significant findings ( $p < 0.05$ )

**Figure 1:**
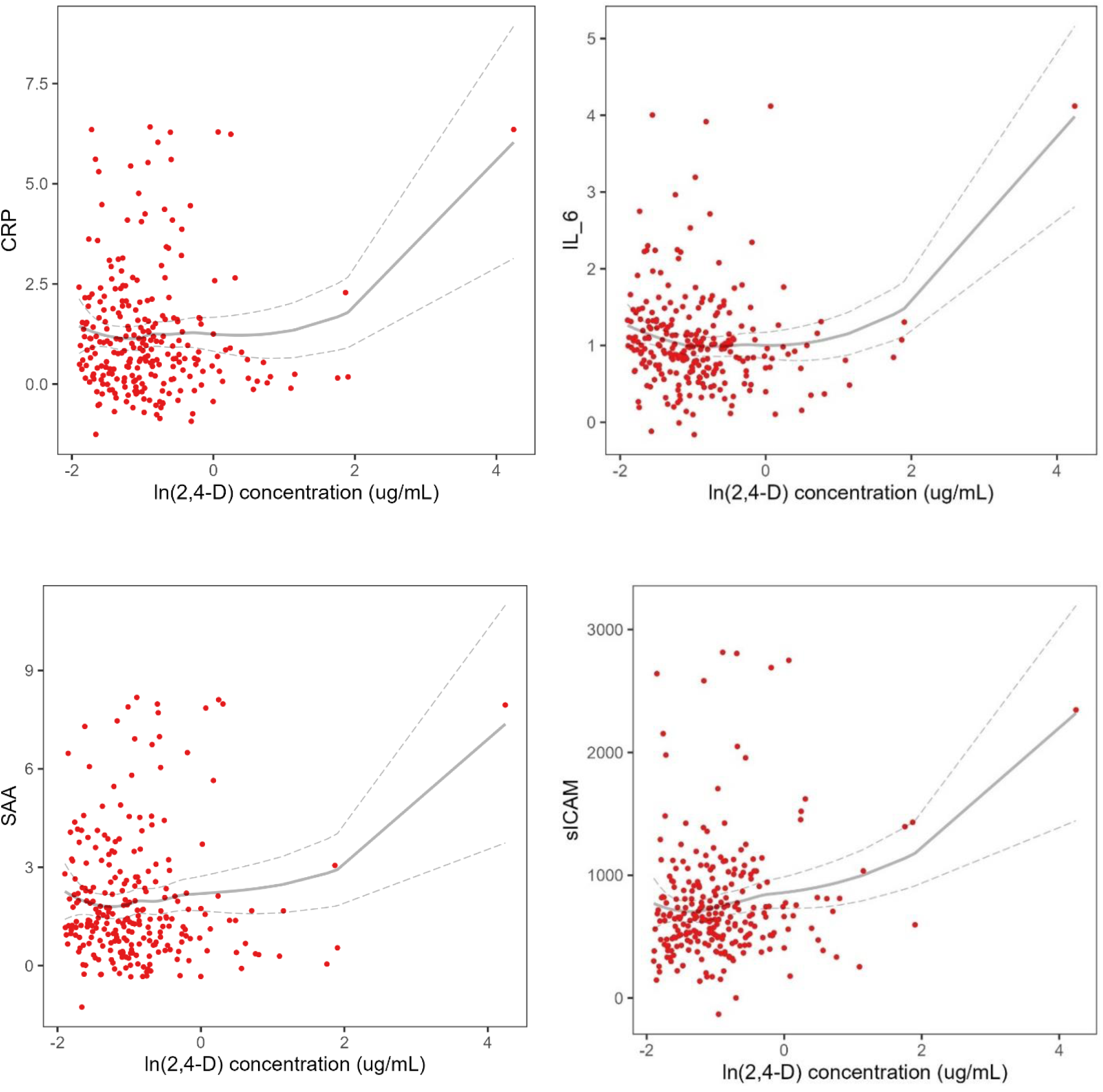
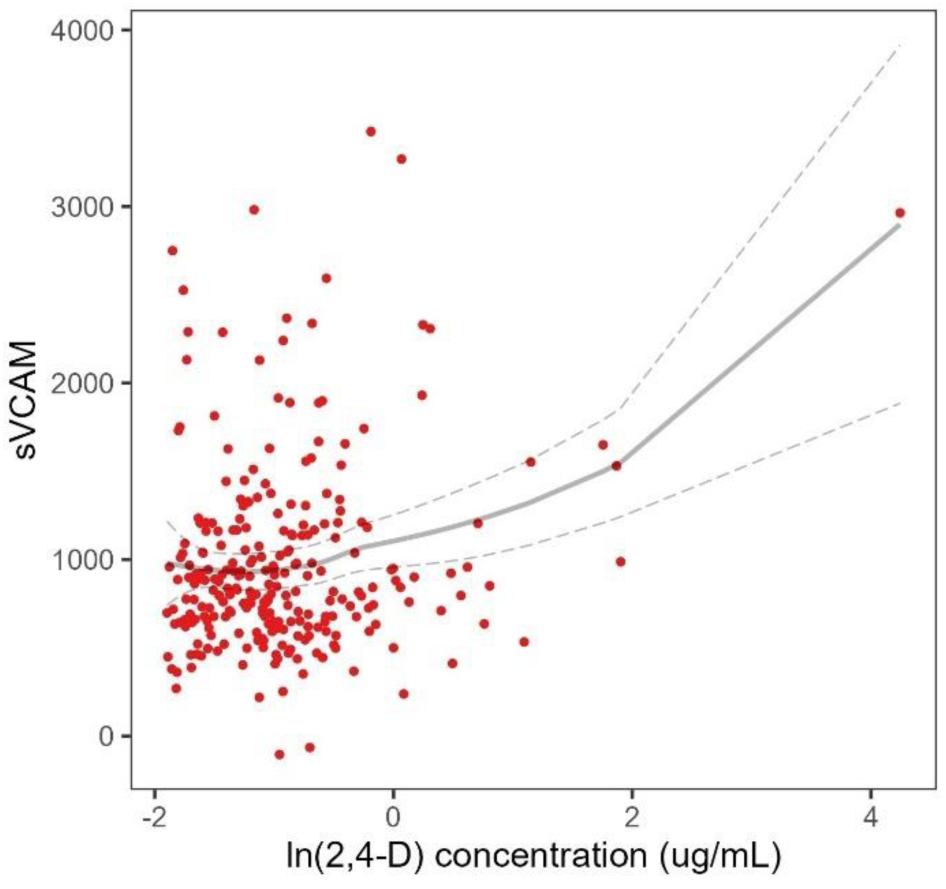
Adjusted associations between pesticide metabolites and inflammatory biomarkers among adolescents, N=507. Red dots represent fully adjusted least squares means for 400 ranks. Gray lines are the LOESS model lines and 95% CI based on the adjusted least squares means data points.

### Associations between Pesticide Metabolite Composites and Inflammatory Markers

Based on the model fit analyses of the PLS components (Table 3), we only included the first composite variable in our models as additional composites did not meaningfully improve the adjusted R-squared value of the model compared with the first composite variable. The loadings of the pesticide metabolite composite 1 are presented in the left panel of Figure 2 in bar graphs. The right panel in Figure 2 depicts the linear model between the first composite variable and each inflammatory biomarker.

**Figure 2.**
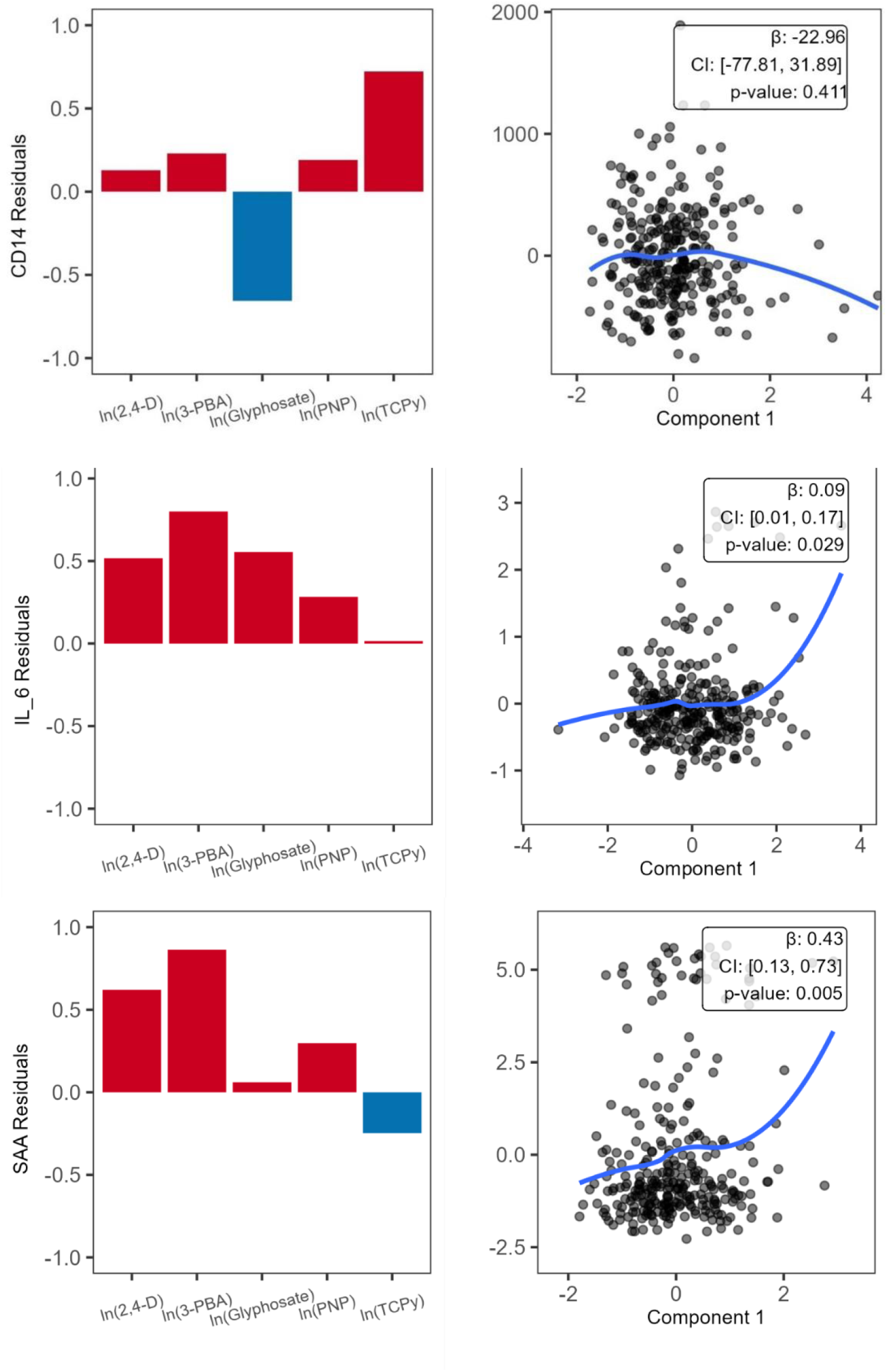
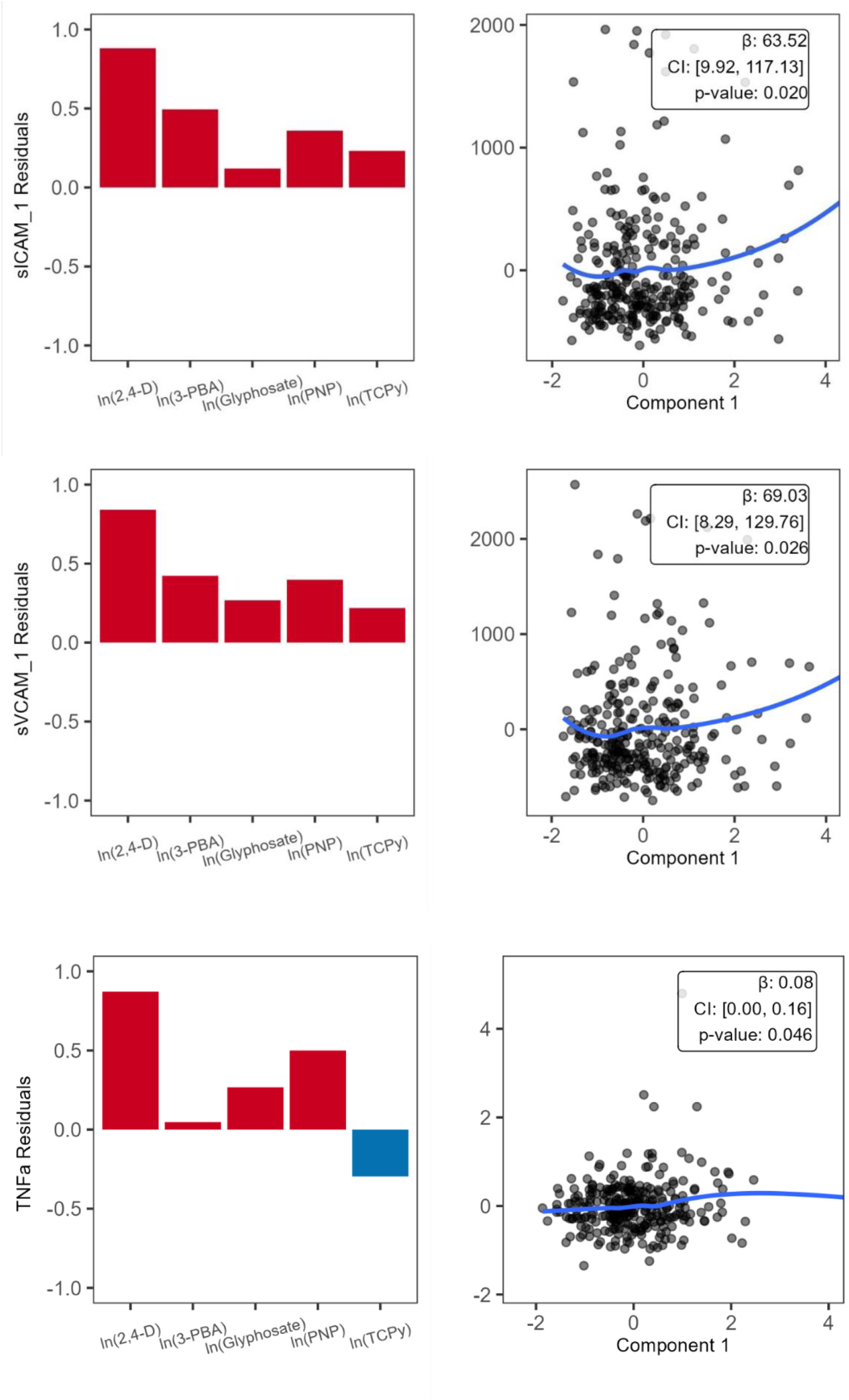
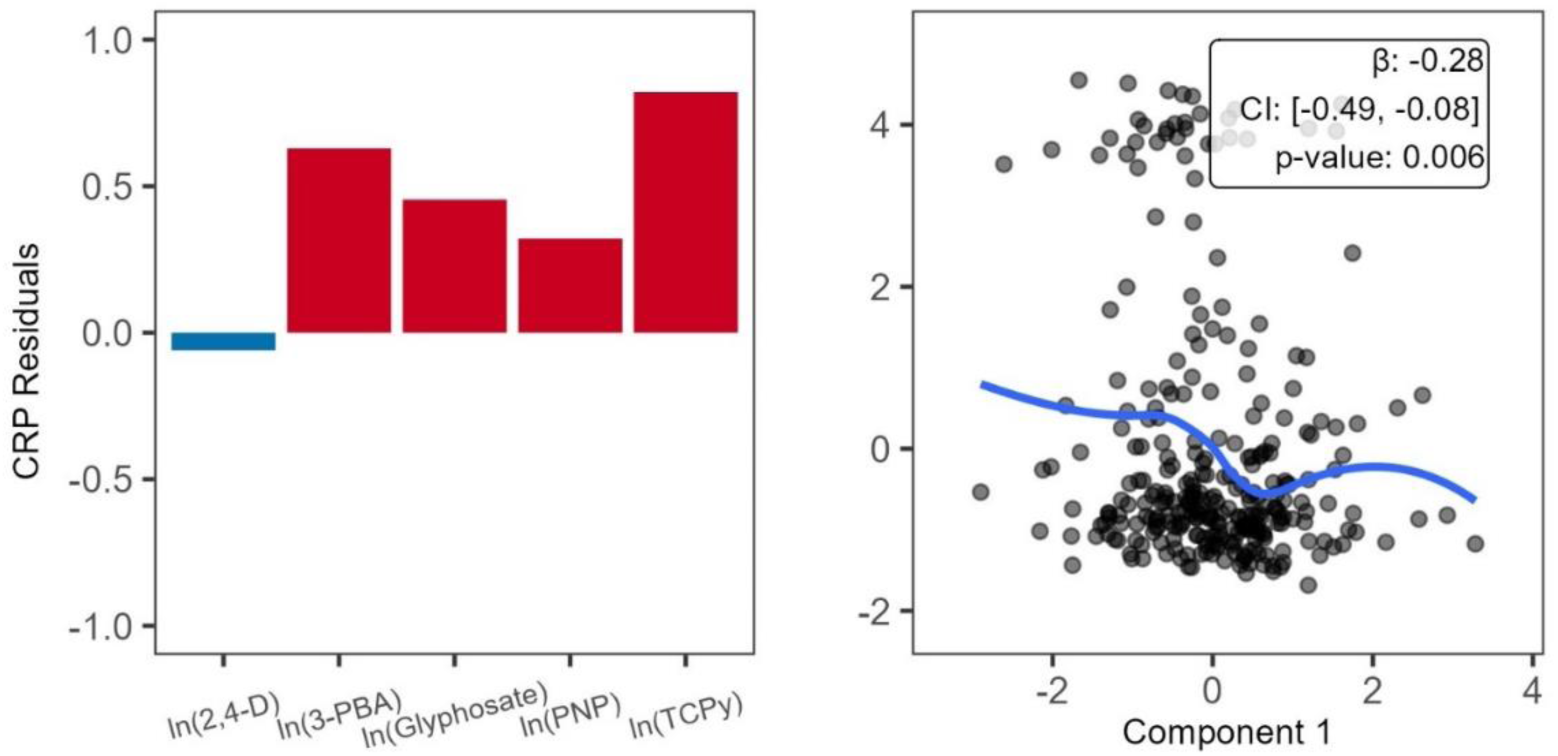
PLS Component 1 loadings and associations between component 1 with inflammation markers in the ESPINA study. N=507 The figures on the left side depict the vector loadings of each independent variable within composite 1. Red color indicates that the component contributes to the same direction as the composite, and blue indicates that it contributes to the opposite direction. The figures on the right are estimated curves depicting the relationship between the composite variable and each inflammatory biomarker with β-coefficient estimates, confidence intervals (CI) and p-values shown in the top left-hand corner of each graph. β represents percent difference in inflammatory marker concentration for a 10% increase in urinary metabolite concentration

**Table 3.** Associations using PLS regression for the associations of pesticide and DEET metabolites with plasma inflammatory markers in adolescents, N= 507.

| Inflammatory Marker | Full model | PLS |  |
| --- | --- | --- | --- |
|  |  | Component 1 | Component 1 +<br>Component 2 |
| sCD-14 | 0<sup>1</sup> | 0.001 | 0<sup>1</sup> |
| CRP | 0.017 | 0.023 | 0.027 |
| IL-6 | 0.003 | 0.013 | 0.013 |
| SAA | 0.014 | 0.024 | 0.024 |
| sICAM-1 | 0.006 | 0.015 | 0.016 |
| sVCAM-1 | 0.003 | 0.014 | 0.013 |
| TNF-α | 0<sup>1</sup> | 0.010 | 0.008 |

Displayed values are adjusted $R^2$ for 3 models: full model, which includes the linear combination of the pesticide variables, including the standardized pesticide variables in one model (2,4-D, Glyphosate, TCPy, 3-PBA, PNP), PLS model including the 1<sup>st</sup> component only and PLS model including the 1<sup>st</sup> and 2<sup>nd</sup> components. The outcomes (inflammatory markers CD-14, CRP, IL-6, SAA, sICAM-1, sVCAM-1, TNF-α and ETU) are residualized for the following covariates: age, gender, race, height-for-age z-score, and parental education. Bolded text indicates statistical significance for 95% confidence.
<sup>1</sup> Negative adjusted $R^2$ values were set to 0

Overall, there were consistent positive associations between pesticide composite 1 and IL-6 (β= 0.09, [0.01. 0.17]), SAA (β= 0.43, [0.13, 0.17]), sICAM-1 (β= 63.52, [9.92, 117.13]), sVCAM-1 (β= 69.03, [8.29, 129.76]) and TNF-*α* (β= 0.08, [0.00, 0.16]), whereas a negative association was observed with CRP (β= -0.28, [-0.49, -0.08], Figure 2), where β represents an increase in the inflammatory biomarker concentration for every unit increase in the composite pesticide variable. Of note, the loadings of the composite variable showed consistent positive associations between 2,4-D, 3-PBA and PNP and the inflammatory biomarkers under study with significant associations. The only exception is for 2,4-D for the association between the composite pesticide variable and CRP.

## Discussion

Our investigation reveals that urinary pesticide metabolite concentrations in adolescents residing in agricultural regions of Ecuador are indeed correlated with heightened levels of inflammatory biomarkers in plasma, aligning with our initial hypothesis. The cumulative exposure to pesticides demonstrates significant positive associations with the blood concentrations of SAA, sVCAM-1, and TNF-*α*, while exhibiting a negative association with CRP. Upon conducting individual analyses of pesticides, it became evident that quadratic associations more effectively elucidate the intricate relationship between several metabolites and biomarkers, indicating non-linear interdependencies.

In particular, concentrations of 2,4-D showed positive quadratic associations with multiple inflammatory biomarkers, including CRP, IL-6, SAA, sICAM-1 and sVCAM-1. These findings are consistent with in-vitro and animal studies indicating 2,4-D can act as an oxidative stressor and immune-activator (Bukowska 2003; Mountassif et al. 2008; Zhou et al. 2022). Proposed mechanisms of how 2,4-D might increase inflammation have been gleaned from in-vitro studies in cell models. Zhou et al. have shown that 2,4-D increases radical oxygen species (ROS) in cells and can directly activate the NOD-, LRR- and pyrin domain-containing protein 3 (NLRP3) inflammasome, which leads to caspase-1 dependent activation of the proinflammatory cytokines IL-1B and IL-18 (Zhou et al. 2022). The evidence that 2,4-D is an oxidative stressor is corroborated by a study by Bukowaska, which showed that 2,4-D depleted the reduced glutathione stores of erythrocytes in vitro (Bukowska 2003). Oxidative stress can lead to inflammatory responses, as cellular pattern recognition receptors (PRRs) detect oxidative danger-associated molecular patterns (DAMPs) (Mittal et al. 2014). Redox-sensitive transcription factors such as NF-kB and AP-1 also directly upregulate inflammatory mediators such as ICAM-1 in response to radical oxygen species (Mittal et al. 2014). Furthermore, IL-1B and IL-18 released through inflammasome activation have both been implicated in the upregulation of ICAM-1 and VCAM-1, suggesting that these cytokines mediate the recruitment of leukocytes into tissues (Meager 1999; Morel et al. 2001). Chronic low-level exposure to 2,4-D may therefore maintain increased oxidative stress and therefore a proinflammatory state in exposed people.

Using PLS analysis, we also found that the composite pesticide metabolite variable was positively associated with IL-6, SAA, sICAM-1, sVCAM-1 and TNF-*α*, and negatively associated with CRP. The PLS allows us to study how independent variables that exhibit collinearity explain changes in the dependent variable and how multiple exposures might synergize in their effect on an outcome. Pesticides are commonly applied in mixtures versus using singular chemicals, increasing the likelihood that agricultural communities will be exposed to a variety of pesticide chemicals simultaneously (Hernández et al. 2013). Thus, it is expected that different urinary pesticide metabolites would be correlated. The ESPINA study participants reported non-engagement in agricultural activities, indicating presumptive low-dose exposure to pesticides via multiple routes, including pesticide drift from nearby agricultural sites to residential areas, parental "take-home" routes from occupational exposure, dietary intake, and other ambient environmental sources (Curl et al. 2002; Coronado et al. 2006). A review of the pesticides utilized in global floriculture identified a total of 201 distinct pesticides, with prevalent classes including organophosphates, carbamates, triazoles, and pyrethroids (Pereira et al. 2021). Hence, mixture modeling using PLS allows us to study interactions, synergies or potentiating effects of pesticides in tandem.

Our findings revealed positive associations between the pesticide composite 1 with levels of IL-6 and TNF-*α*, which are used as general markers of inflammation. IL-6 and TNF-*α* are principal cytokines with pleiotropic functions that regulate inflammatory state and several key physiologic processes (Kalliolias and Ivashkiv 2016). The immune roles of the IL-6 and soluble IL-6 receptor (sIL-6R) complex include the progression from acute to chronic inflammatory states and the engagement of the adaptive immune system following innate response. TNF-*α*’s pro-inflammatory functions include upregulating selectins, CAMs, and pro-inflammatory NF-KB (Zelová and Hošek 2013). Animal and in vitro studies have found that perinatal exposure to glyphosate increased the levels of IL-6 and TNF-*α* in rat livers and upregulated IL-6 and TNF-*α* mRNA and protein expression (Bai et al. 2022; Rieg et al. 2022). Furthermore, the mRNA and protein expression of both IL-6 and TNF-*α* are broadly upregulated with various classes of pesticides, including neonicotinoids, pyrethroids, organophosphates, and herbicides (Mense et al. 2006; Agrawal et al. 2015a; Hossain et al. 2017; Pandey et al. 2019; Miao et al. 2022; Deng et al. 2022).

Our results revealed associations between pesticide exposure with soluble levels of multiple vascular inflammatory markers, namely cellular adhesion molecules (CAMs) and the acute phase reactant SAA, both of which are also regulated by IL-6 and TNF-*α*. The aforementioned IL-6 and sIL-6R complex facilitates the expression of cellular adhesion molecules (CAMs) on endothelial cells and is involved in T-cell differentiation (Scheller et al. 2011). CAMs allow for the extravasation of monocytes and lymphocytes into tissues and are therefore key inflammatory molecules. High levels of CAMs are involved in the pathogenesis of debilitating diseases such as atherosclerosis (Yin et al. 2021). Like CAMs, SAA may play a role in the development of atherosclerosis by promoting increased release of vascular proteoglycans, which bind to and retain lipids in the endothelial wall through ionic interactions (Wilson et al. 2008; Thompson et al. 2015). Notably, the herbicides 2,4-D (primarily) and glyphosate had consistent positive quadratic associations with sVCAM-1 and/or sICAM-1, evincing a potential role of herbicides in promoting the underlying pathophysiological processes by which atherosclerosis and neuroinflammation might occur. Epidemiological evidence from other studies has demonstrated that pesticide exposure is a risk factor for atherosclerosis. One cross-sectional study of 447 middle-aged and older participants in a rural area of Korea revealed a significantly greater atherosclerotic risk associated with reported pesticide exposure (Park et al. 2020). Another epidemiological study reported a positive association between cardiovascular disease incidence and self-reported pesticide exposure in 7,557 middle-aged Japanese American men living in Hawaii (Berg et al. 2019). Together, our findings indicate that the associations between pesticide exposure and general and vascular inflammation may underlie inflammatory disease outcomes reported among individuals with chronic pesticide exposure.

The observed negative associations between pesticide exposure and CRP concentrations are intriguing but unexplained, especially considering the positive correlations noted between CRP concentrations and other measured inflammation markers. Given that CRP exists in two isoforms with potentially opposing roles such that a soluble pentameric form found in circulating blood may be involved in the resolution of inflammation compared to a monomeric form which is localized in inflamed tissues (Del Giudice and Gangestad 2018), our observation may reflect a complex phenomenon. Nonetheless, the participants in this study were children and adolescents with no known inflammatory conditions, in whom the levels of CRP are generally low and its implications for health outcomes are not well established other than small to modest associations with some demographic factors and adiposity (Cook et al. 2000; Caballero et al. 2008; Nappo et al. 2013). Investigations of CRP levels in children remain scarce besides a few studies of CRP as a clinical test for acute infections (Staiano et al. 2023) or inflammatory conditions (Enocsson et al. 2021), and its baseline levels in children in the absence of inflammatory diseases or infections are often low which is also shown in our study participants (i.e., mean 1.4 µg/mL) (Schlenz et al. 2014). Thus, it is premature to speculate on the health or mechanistic implications of this finding.

Understanding the associations between urinary metabolites and inflammatory biomarkers is important in this population given that they have a higher exposure to pesticides. Creatinine-corrected pesticide metabolite concentrations (ug/L) in the ESPINA cohort were compared to creatinine-corrected concentrations reported by the National Health and Nutrition Examination Survey (NHANES). The geometric means of urinary metabolite concentrations from the 2015–2016 NHANES 12–19-year-old study participants were referenced (National Health and Nutrition Examination Survey 2016). Geometric means of the creatinine-corrected urinary metabolite concentrations for the ESPINA cohort were calculated and compared to geometric means of the 2015–2016 NHANES cohort. DEET metabolite concentrations were higher in the NHANES population (Table S 2). However, all the neonicotinoid concentrations and three of the four organophosphate metabolites were higher in the ESPINA participants. Overall, the ESPINA population had higher concentrations of neonicotinoid and organophosphate metabolites in their urine as compared to the participants in the United States NHANES study (AND 0.31 vs 0.18; CLOT 0.17 vs 0.12; IMID 0.34 vs 0.23; 5-Hydroxy imidacloprid 0.48 vs 0.28; 3,5,6-Trichloro-2-pyridinol 3.07 vs 0.92; MDA 3.07 vs 0.92; 2-isopropyl-4-methyl-6-hydroxypy 0.11 vs 0.07). However, there were lower levels of DCBA, ECBA and acetamiprid metabolites, and the same levels of PNP and 2,4-D when comparing ESPINA participants to those in the NHANES study (DCBA 0.63 vs 4.42; ECBA 0.36 vs 1.77).

Our study stands out due to its comprehensive examination of a diverse panel of pesticides, including insecticides, herbicides, and insect repellents, exploring both their individual effects and their combined associations with markers of overall and vascular inflammation. Additionally, our investigation focuses on adolescents, a demographic lacking sufficient evidence in similar studies, particularly those residing in rural areas of Ecuador, which are often underrepresented in research endeavors. Another strength of this study is the use of PLS regression analysis, which allows for the integration of the synergistic effects of multiple pesticides on inflammation. The PLS analyses also allow us to investigate the relationships between collinear pesticide metabolites with the inflammatory biomarker outcomes. Finally, the analysis in our study also does not rely on reported pesticide exposure but rather uses urinary pesticide metabolites to measure the level of exposure quantitatively.

A limitation of this study lies in the exclusive examination of circulating inflammatory biomarkers, which may not fully reflect tissue- and organ-specific inflammatory alterations. Furthermore, the cross-sectional nature of our analyses precludes definitive determination of causality and, in theory, the directionality of the association (do urinary pesticide concentrations influence inflammation markers? Or do inflammation markers influence urinary pesticide concentrations?). We are unaware of mechanisms for how circulating inflammatory markers could influence urinary pesticide concentrations, although this is theoretically possible if pesticide metabolism and excretion processes are modified by circulating inflammatory markers. However, although it seems more likely that pesticides may influence the concentrations of inflammatory markers as observed in the experimental studies described in this manuscript, it is plausible that inflammatory markers may modulate the metabolism and excretion of pesticides. This topic needs further evaluation. Future studies of inflammatory-disease incidence in subjects chronically exposed to pesticides could help reveal any clinical manifestations of changes in inflammation status secondary to pesticide exposure. More prospective studies examining how inflammation status changes in response to low-grade chronic exposure to pesticides could help reveal more about the nature of physiological responses to agrochemicals.

## Conclusion

We found that overall pesticide exposure, estimated using a pesticide composite variable of urinary pesticide metabolites, was positively associated with IL-6, SAA, sICAM-1, sVCAM-1 and TNF-*α*, and negatively with CRP. Our findings also indicate that some of these associations are likely non-linear in nature, indicated by the consistent curvilinear associations of the herbicide 2,4-D with CRP, IL-6, SAA, sICAM-1 and sVCAM-1. These results agree with existing in vitro and animal studies that describe the proinflammatory nature of pesticides in tissue culture and animal models. Our study is one of the first to assess the associations between pesticide exposure, measured using urinary pesticide metabolites, and serum inflammation markers in adolescents and children. Our results suggest that exposure to pesticides at levels lower than those that can cause acute toxicity may alter the inflammatory state in the human body, which may be a mechanism by which pesticide exposure leads to detrimental health effects. Given the potential role of pesticides in inflammation, attention should be given to the incidence and prevalence of inflammatory diseases in affected populations. Given the cross-sectional design of this study, replication through longitudinal analysis would offer a more comprehensive understanding of the causal pathway linking pesticide exposure to inflammation. These findings underscore the imperative to reassess the widespread use of pesticides in agriculture, emphasizing the importance of adopting sustainable and health-conscious approaches to agricultural practices.

## Data Availability

All data produced in the present study are available upon reasonable request to the authors

https://knit.ucsd.edu/espina/

## Acknowledgments

We especially thank all the participants who participated in the present study. We thank the ESPINA study staff, Fundación Cimas del Ecuador, the Parish Governments of Pedro Moncayo County, the community members of Pedro Moncayo and the Education District of Pichincha-Cayambe-Pedro Moncayo counties for their contributions to and support of this project. The coauthors extend thanks to Dolores Lopez Paredes for her contributions to this research.

## Author Contributions

Conceptualization, Funding, & Methodology: **SH, JST & JRS** – Fieldwork, Data Collection & Management: **DM, BNC, JST & JRS** – Data Analysis & Interpretation**: KY, XT, JRS & SH** – Original Draft Preparation: **MNH** – Literature Review, Manuscript Writing – Review & Editing: **MNH, BNC, XT, RPP, JST, DB, SH & JRS**.

All the coauthors read and approved the final manuscript.

## Consent to participate and Ethics

Informed consent, parental authorization of child participation and assent of child participants older than 7 years of age were obtained. For the 2016 examination of the ESPINA study, we obtained approval from the Institutional Review Boards of Boards of the University of California San Diego (UCSD), Universidad San Francisco de Quito and Ecuador’s Ministry of Public Health in 2016.

---

**Table S1.** Correlations between inflammatory biomarkers.

|  | **CD14** | **CRP** | **IL-6** | **SAA** | **sICAM-1** | **sVCAM-1** | **TNF-α** |
| --- | --- | --- | --- | --- | --- | --- | --- |
| **CD14** | 1 | 0.06 | 0.02 | **0.09** | 0 | 0.02 | **0.24** |
| **CRP** |  | 1 | 0.05 | **0.63** | **0.52** | **0.51** | 0.08 |
| **IL-6** |  |  | 1 | 0.03 | 0.01 | 0.01 | 0.04 |
| **SAA** |  |  |  | 1 | **0.4** | **0.46** | 0.03 |
| **sICAM-1** |  |  |  |  | 1 | **0.88** | **0.22** |
| **sVCAM-1** |  |  |  |  |  | 1 | **0.25** |
| **TNF-α** |  |  |  |  |  |  | 1 |

Values in bold were significant at a $p < 0.05$ level

**Table S2.**
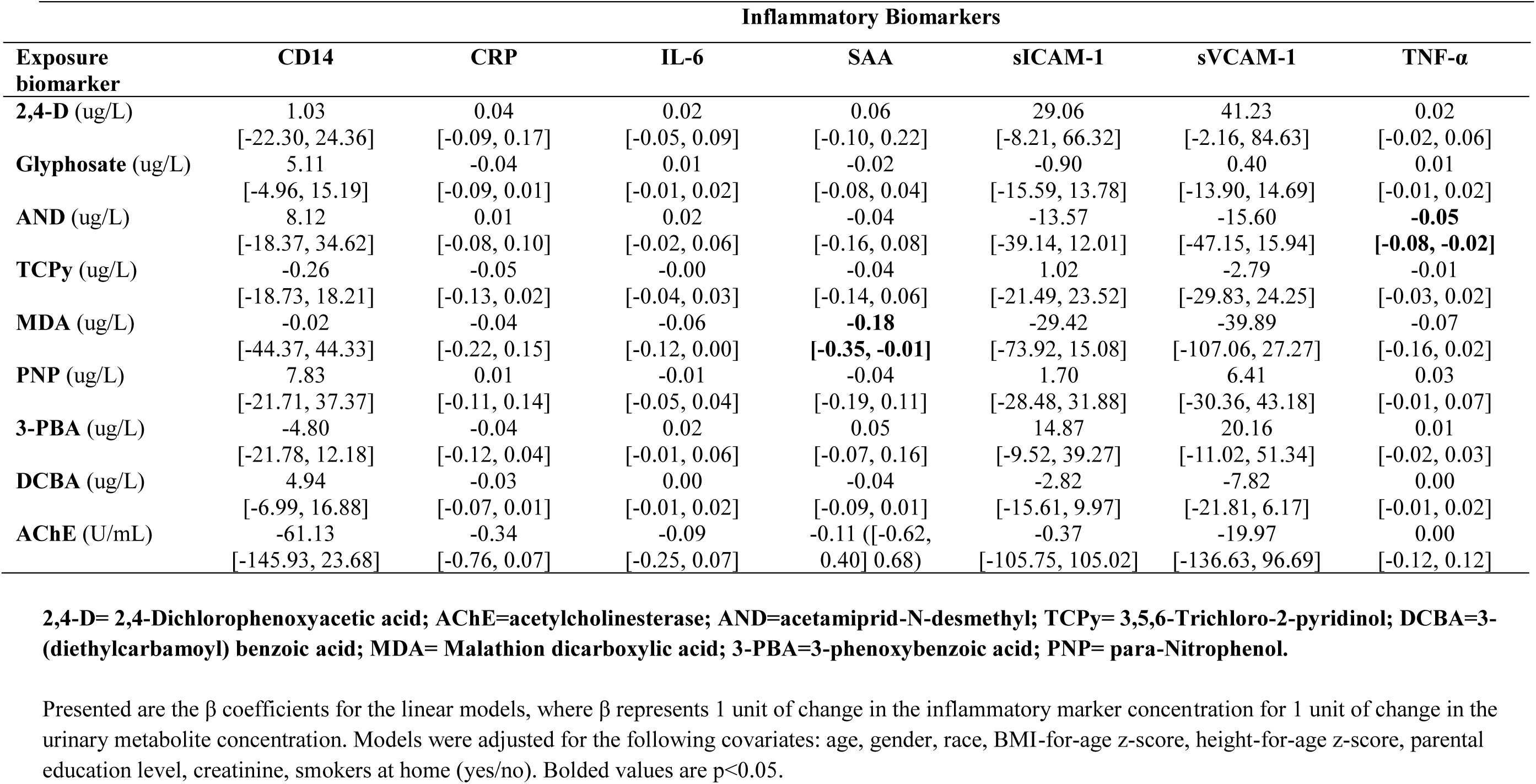
Independent linear associations between biomarkers of pesticide exposure and inflammation. N=507.

| Exposure biomarker | Inflammatory Biomarkers |  |  |  |  |  |  |
| --- | --- | --- | --- | --- | --- | --- | --- |
| | CD14 | CRP | IL-6 | SAA | sICAM-1 | sVCAM-1 | TNF-α |
| **2,4-D** (ug/L) | 1.03<br>[-22.30, 24.36] | 0.04<br>[-0.09, 0.17] | 0.02<br>[-0.05, 0.09] | 0.06<br>[-0.10, 0.22] | 29.06<br>[-8.21, 66.32] | 41.23<br>[-2.16, 84.63] | 0.02<br>[-0.02, 0.06] |
| **Glyphosate** (ug/L) | 5.11<br>[-4.96, 15.19] | -0.04<br>[-0.09, 0.01] | 0.01<br>[-0.01, 0.02] | -0.02<br>[-0.08, 0.04] | -0.90<br>[-15.59, 13.78] | 0.40<br>[-13.90, 14.69] | 0.01<br>[-0.01, 0.02] |
| **AND** (ug/L) | 8.12<br>[-18.37, 34.62] | 0.01<br>[-0.08, 0.10] | 0.02<br>[-0.02, 0.06] | -0.04<br>[-0.16, 0.08] | -13.57<br>[-39.14, 12.01] | -15.60<br>[-47.15, 15.94] | **-0.05**<br>**[-0.08, -0.02]** |
| **TCPy** (ug/L) | -0.26<br>[-18.73, 18.21] | -0.05<br>[-0.13, 0.02] | -0.00<br>[-0.04, 0.03] | -0.04<br>[-0.14, 0.06] | 1.02<br>[-21.49, 23.52] | -2.79<br>[-29.83, 24.25] | -0.01<br>[-0.03, 0.02] |
| **MDA** (ug/L) | -0.02<br>[-44.37, 44.33] | -0.04<br>[-0.22, 0.15] | -0.06<br>[-0.12, 0.00] | **-0.18**<br>**[-0.35, -0.01]** | -29.42<br>[-73.92, 15.08] | -39.89<br>[-107.06, 27.27] | -0.07<br>[-0.16, 0.02] |
| **PNP** (ug/L) | 7.83<br>[-21.71, 37.37] | 0.01<br>[-0.11, 0.14] | -0.01<br>[-0.05, 0.04] | -0.04<br>[-0.19, 0.11] | 1.70<br>[-28.48, 31.88] | 6.41<br>[-30.36, 43.18] | 0.03<br>[-0.01, 0.07] |
| **3-PBA** (ug/L) | -4.80<br>[-21.78, 12.18] | -0.04<br>[-0.12, 0.04] | 0.02<br>[-0.01, 0.06] | 0.05<br>[-0.07, 0.16] | 14.87<br>[-9.52, 39.27] | 20.16<br>[-11.02, 51.34] | 0.01<br>[-0.02, 0.03] |
| **DCBA** (ug/L) | 4.94<br>[-6.99, 16.88] | -0.03<br>[-0.07, 0.01] | 0.00<br>[-0.01, 0.02] | -0.04<br>[-0.09, 0.01] | -2.82<br>[-15.61, 9.97] | -7.82<br>[-21.81, 6.17] | 0.00<br>[-0.01, 0.02] |
| **AChE** (U/mL) | -61.13<br>[-145.93, 23.68] | -0.34<br>[-0.76, 0.07] | -0.09<br>[-0.25, 0.07] | -0.11 ([-0.62, 0.40] 0.68) | -0.37<br>[-105.75, 105.02] | -19.97<br>[-136.63, 96.69] | 0.00<br>[-0.12, 0.12] |

**2,4-D= 2,4-Dichlorophenoxyacetic acid; AChE=acetylcholinesterase; AND=acetamiprid-N-desmethyl; TCPy= 3,5,6-Trichloro-2-pyridinol; DCBA=3-(diethylcarbamoyl) benzoic acid; MDA= Malathion dicarboxylic acid; 3-PBA=3-phenoxybenzoic acid; PNP= para-Nitrophenol.**
Presented are the $\beta$ coefficients for the linear models, where $\beta$ represents 1 unit of change in the inflammatory marker concentration for 1 unit of change in the urinary metabolite concentration. Models were adjusted for the following covariates: age, gender, race, BMI-for-age z-score, height-for-age z-score, parental education level, creatinine, smokers at home (yes/no). Bolded values are $p < 0.05$ .

